# Shared genetic architecture and causal pathways between attention deficit hyperactivity disorder and restless legs syndrome

**DOI:** 10.1101/2024.06.20.24309235

**Authors:** Fu-Jia Li, Jin-Yu Li, Ru-Yu Zhang, Xuan-Jing Liu, Bing-Chen Lv, Tao Zhang, Yu-Ning Liu, Zi-Xuan Zhang, Wei Zhang, Gui-Yun Cui, Chuan-Ying Xu

## Abstract

Previous studies have revealed a significant overlap between ADHD and RLS populations, with shared pathological mechanisms such as dopaminergic function and iron metabolism deficits. However, the genetic mechanisms underlying these connections remain unclear. In our study, we conducted a genome-wide genetic correlation analysis to confirm a shared genetic structure between ADHD and RLS. We identified five pleiotropic loci through PLACO analysis, with colocalization analysis revealing a shared causal genetic variant, rs12336113, located in an intron of the PTPRD gene within one of these loci. Additionally, we identified 14 potential shared genes and biological pathways between these diseases. Protein-protein interaction analysis demonstrated close interactions among six genes: PTPRD, MEIS1, MAP2K5, SKOR1, BTBD9, and TOX3. We further investigated gene-driven causal pathways using univariable Mendelian randomization (MR), multivariable MR, and Network MR analyses. Our findings indicate that ADHD may indirectly promote the onset of RLS by advancing the age of first birth, while RLS could indirectly contribute to ADHD by reducing fractional anisotropy in body of corpus callosum. Notably, an increase in radial diffusivity, rather than a decrease in axial diffusivity, played a crucial role in this process. In conclusion, our research supports a close genetic link between ADHD and RLS, identifying PTPRD as the most likely pleiotropic gene between these conditions. Moreover, ADHD may indirectly promote RLS onset by advancing the age of first birth, while RLS may indirectly promote ADHD onset by causing demyelination in body of corpus callosum.

## Introduction

Attention-deficit/hyperactivity disorder (ADHD) is a neurodevelopmental disorder characterized by persistent inattention, hyperactivity, and impulsivity^1^. It is estimated that 5-7% of school-age children have ADHD^2,3^, and up to 70% of those with childhood-onset ADHD continue to exhibit symptoms into adulthood^4^. ADHD is associated with risk-taking behaviors and various neuropsychiatric conditions, significantly increasing the risk of developing chronic diseases in the future. Consequently, this disease imposes a substantial societal burden^5^.

Restless legs syndrome (RLS) is a common sensorimotor disorder with a prevalence of 6%-12%^6^. It is characterized by intensely unpleasant sensations and an almost irresistible urge to move the legs in the evening or at night^7^. Temporary symptomatic relief may be achieved through leg movement or walking, significantly impacting sleep quality and overall quality of life for patients.

The comorbidity of ADHD and RLS is an intriguing phenomenon. Observational studies have shown a significant population overlap between ADHD and RLS. Patients with ADHD, whether children, adolescents, or adults, are more likely to exhibit RLS symptoms compared to healthy individuals^8–10^. Conversely, individuals with RLS, regardless of age, have a higher prevalence of ADHD, with more severe symptoms^11–13^. Additionally, cross-generational and family history studies suggest a significant genetic link between ADHD and RLS. Parents of children with ADHD have a higher incidence of RLS^14,15^, and children with a family history of RLS exhibit more severe ADHD symptoms^12^.

Several explanations have been proposed for the comorbidity. One hypothesis suggests that RLS may lead to ADHD symptoms by causing sleep fragmentation and reduced sleep quality^16^. Another theory posits that ADHD and RLS share common pathologies, such as dopaminergic dysfunction in the central nervous system (CNS)^17–19^ and iron metabolism disorders, including reduced serum iron levels^20–22^. Notably, these two pathological changes are closely linked. Iron is a cofactor for tyrosine hydroxylase, the rate-limiting enzyme in dopamine synthesis. Iron metabolism disorders have been shown to alter the density and activity of dopamine D1 and D2 receptors, as well as the dopamine transporter (DAT)^23,24^.

The potential genetic link and shared pathological mechanisms between ADHD and RLS prompted us to investigate whether there are common genetic variants underlying both disorders. In 2009, B.G. Schimmelmann and his team conducted a genome-wide association study (GWAS) involving 224 families (including 386 children with ADHD and their parents)^25^. The study found no significant association between single nucleotide polymorphisms (SNPs) in known RLS risk genes (MEIS1, BTBD9, and MAP2K5) and ADHD. We hypothesize that the small sample size and limited target genes may have hindered the detection of potential positive results.

With advancements in GWAS techniques and access to higher-quality data, we had the opportunity to explore the genetic mechanisms underlying the comorbidity of ADHD and RLS more thoroughly. This study proceeded in three stages. In **stage one**, we performed a genome-wide genetic correlation analysis to determine whether ADHD and RLS share a common genetic structure, which can result from pleiotropy (where a genetic variant affects both traits) and/or causality (where a genetic variant affects a trait via its effect on an intermediate trait). In **stage two**, using cross-trait pleiotropic analysis under composite null hypothesis (PLACO) and systematic downstream analyses, we identified pleiotropic loci and shared genes between ADHD and RLS, focusing on those related to iron metabolism and dopaminergic neuron function. In **stage three**, Mendelian Randomization (MR) analysis leveraged its advantage of being less susceptible to confounding bias and reverse causation, allowing us to effectively explore the causal relationship between ADHD and RLS. Additionally, Network MR analysis further elucidated indirect causal pathways between these two diseases, such as whether RLS can lead to ADHD by inducing sleep disturbances.

## Methods

### 1 Data characteristics

This study is a secondary analysis of existing GWAS data, all of which were publicly available with no original data collection involved. Each included study received approval from their respective institutional ethics review committees, and informed consent was obtained from all participants. We selected data based on the following criteria: 1) European populations, 2) the largest publicly available datasets, and 3) minimal sample overlap in MR analysis. Detailed characteristics of the data used in this study are described in **Table S1**.

#### 1.1 ADHD

The GWAS data for ADHD were derived from three independent cohorts, totaling 225,543 participants (38,691 cases and 186,843 controls)^26^. Specifically, participants from iPSYCH included 25,895 cases and 37,148 controls, with ADHD diagnosed by psychiatrists according to ICD-10 criteria (F90.0, F90.1, F98.8)^27^. Controls were randomly selected from the same nationwide birth cohort and did not have an ADHD diagnosis. In the deCODE cohort (8,281 cases and 137,993 controls), ADHD cases included individuals with a clinical diagnosis of ADHD (n = 5,583) according to ICD-10 criteria (F90, F90.1, F98.8) or those prescribed medication specific for ADHD symptoms (n = 2,698). The control sample excluded individuals with diagnoses of schizophrenia, bipolar disorder, autism spectrum disorder, or self-reported ADHD symptoms or diagnosis. For the PGC cohort (4,515 cases and 11,702 controls), specific diagnostic criteria can be found in a previous study by Demontis et al^28^.

#### 1.2 RLS

The GWAS data for RLS originated from six independent cohorts, totaling 480,982 participants (10,257 cases and 470,725 controls)^29^. Specifically: (1) The deCODE cohort (2,636 cases and 11,448 controls) assessed RLS using a self-completed questionnaire based on the International RLS Study Group (IRLSSG) diagnostic criteria. (2) Participants in the INTERVAL study (3,065 cases and 24,923 controls) and the DBDS cohort (1,379 cases and 25,186 controls) were assessed using the Cambridge-Hopkins RLS questionnaire (CH-RLSq). (3) In the UK Biobank cohort (1,916 cases and 406,649 controls), RLS status was determined using the ICD-10 code G25.8. (4) In the Donor InSight-III cohort (565 cases and 1,798 controls), RLS status was self-reported using a questionnaire developed as part of the RISE study, based on IRLSSG criteria and created in collaboration with an RLS expert. (5) The Emory cohort (696 cases and 721 controls) had RLS status clinically verified by one of two clinicians and supplemented by objective measurements of periodic leg movements in sleep (PLMS) along with additional secondary and supportive diagnostic features.

#### 1.3 Confounders

Through a systematic literature search, we identified six characteristics that might confound the relationship between ADHD and RLS: body mass index (BMI)^30,31^, smoking behavior^32,33^, serum vitamin D levels^34,35^, estimated glomerular filtration rate (eGFR)^36,37^, serum iron levels^20–22^, and dopaminergic dysfunction in the CNS^17–19^. These factors were adjusted in the MVMR analysis.

Notably, dopaminergic neurons in the CNS primarily function by synthesizing and releasing dopamine, which cannot cross the blood-brain barrier. Unfortunately, we currently lack GWAS data describing dopaminergic dysfunction or cerebrospinal fluid (CSF) dopamine levels. Dopamine 3-O-sulfate is a major metabolite of dopamine, so we used CSF dopamine 3-O-sulfate levels to indirectly reflect dopaminergic function in the CNS^38^.

#### 1.4 Candidate mediators

We explored the bidirectional indirect causal pathways between ADHD and RLS using Network MR analysis. All candidate mediators had to meet the following criteria, supported by previous literature: 1) they may be influenced by the exposure factors, and 2) they may affect the outcome factors. In the causal pathway from ADHD to RLS, we identified five personal behavior traits (smoking behavior, alcohol consumption, use of depression medications, age at first birth, and multiple gestation), two mental illnesses (anxiety disorders and depression), six chronic diseases (obesity, type 2 diabetes mellitus, asthma, ischemic stroke, chronic kidney disease, and migraine), eight iron-sensitive imaging features, and nine DTI features as candidate mediators. Conversely, the pathway from RLS to ADHD included three sleep traits (combined sleep disorders, insomnia, and disorder of the sleep-wake rhythm), eight iron-sensitive imaging features, and nine DTI features. Detailed results of the literature search are provided in **Table S10**.

### 2 Stage one: exploring shared genetic structure

The flowchart of the entire study is shown in **Figure 1**. We used linkage disequilibrium score regression (LDSC, v1.0.01) to estimate the genome-wide genetic correlation between ADHD and RLS^39^, employing European ancestry genotype data from 1000 genomes as the reference panel^40^. High-definition likelihood (HDL) reduces the variance of genetic association estimates by about 60% compared to LDSC^41^. We performed HDL using the R package HDL-v1.4.0, utilizing 1,029,876 well-imputed HapMap3 SNPs as the reference panel.

**Figure 1.**
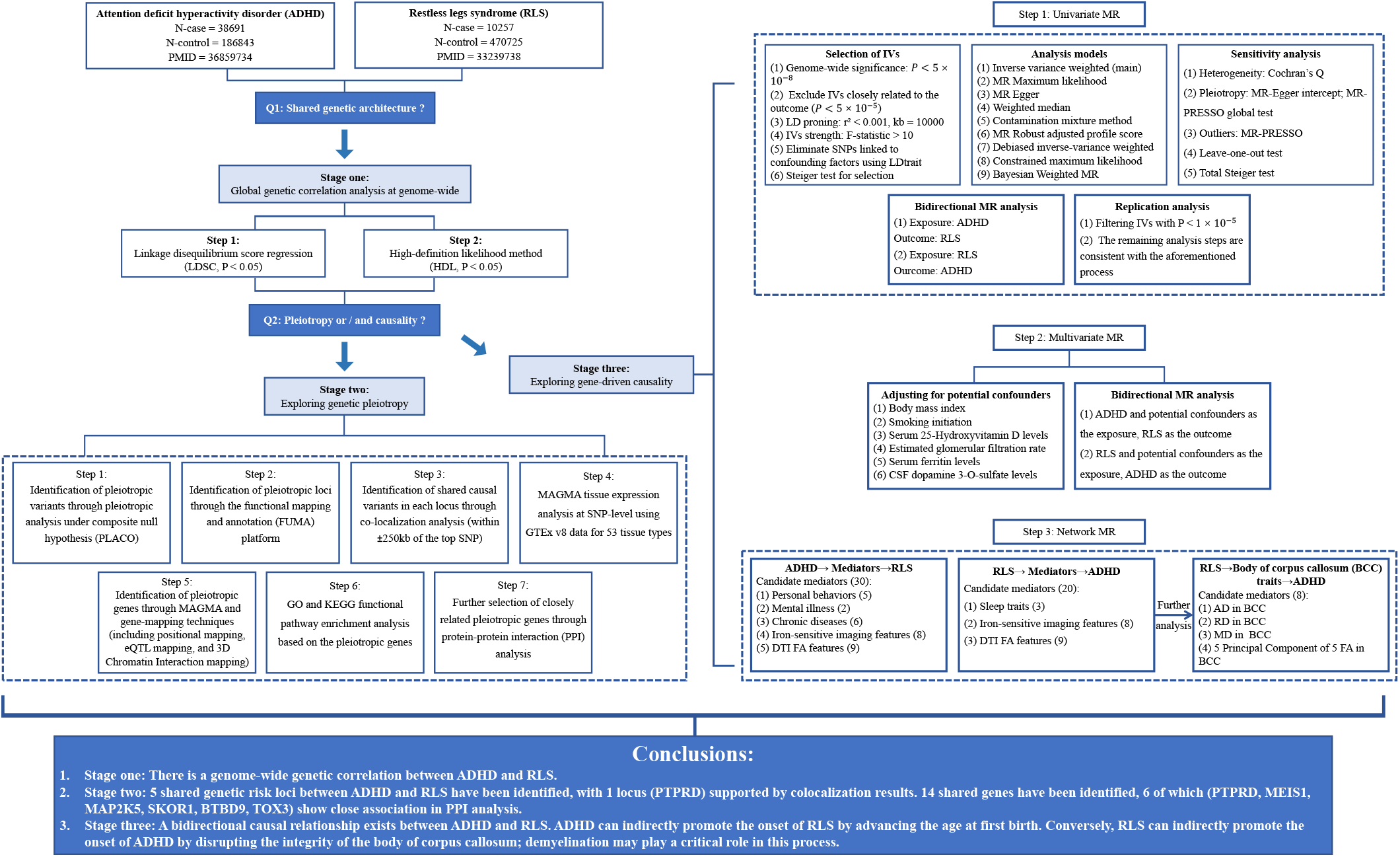
depicts the study design. CSF Cerebrospinal fluid, IVs Instrumental variables, SNP Single nucleotide polymorphism.

### 3 Stage two: exploring genetic pleiotropy

(1) PLACO identified pleiotropic SNPs between ADHD and RLS^42^. (2) Based on PLACO results, functional mapping and annotation (FUMA) characterized pleiotropic loci^43^. (3) Bayesian colocalization analysis using the “coloc” package (v5.2.1) identified shared causal variants in each pleiotropic locus^44^. We analyzed top SNPs within 250kb upstream and downstream of each locus, declaring a colocalized locus with a posterior probability of H4 (PP.H4) greater than 0.75. (4) Tissue enrichment analysis was conducted at the SNP level with 53 available tissue types from GTEx (v.8) using MAGAMA method. (5) We mapped the identified pleiotropic SNPs into shared genes using MAGMA and gene mapping (including positional mapping, eQTL mapping, and 3D Chromatin Interaction mapping). (6) Using the “clusterProfiler” R package, we evaluated the functional enrichment of pleiotropic genes in KEGG^45^ and GO^46^ pathways. (7) We used the STRING (version 12.0) database for protein–protein interaction (PPI) analysis to identify the most closely connected pleiotropic genes^47^. Bonferroni correction was applied for multiple testing in all analyses.

### 4 Stage Three: Exploring gene-driven causality

In this stage, we primarily used the two-sample MR method to investigate the potential causalities between ADHD and RLS. This research adheres to the STROBE-MR guidelines^48^ (**Table S5**).

#### 4.1 Univariable MR

Genetic variants were selected as instrumental variables (IVs) based on strong associations with the exposure (P < 5 × 10^−8^). The R² value of SNPs estimated the variance in exposure, and the F-statistic measured instrument strength. Only SNPs with F-statistic > 10 were included for analysis. Independent SNPs were clumped based on European ancestry reference data (1000 Genomes Project, r² > 0.001, genomic region = 10,000 kb). To meet the independence assumption for MR analysis, 10 IVs for ADHD associated with BMI, serum 25-Hydroxyvitamin D levels, and stroke, and 2 IVs for RLS associated with BMI were excluded (**Table S6**) based on the LDtrait query (https://ldlink.nih.gov/?tab=ldtrait).

The Inverse-Variance Weighted (IVW) method^49^ was our primary analysis due to its accuracy and power when all selected SNPs are valid IVs. To ensure robustness, we applied eight secondary MR methods: Maximum Likelihood (ML)^50^, Weighted Median (WM)^51^, MR-Egger regression^52^, Constrained Maximum Likelihood - Model Average (cML-MA)^50^, contamination mixture method (ConMix)^53^, robust adjusted profile score (MR-RAPS) ^54^, debiased IVW (dIVW), and the Bayesian Weighted MR (BWMR)^55^.

**To clarify the direction of causality**, we implemented the following measures: (1) excluded IVs significantly associated with the outcome (P < 5 × 10^−5^) ^56^, (2) further filtered IVs using the Steiger filtration^57^, (3) performed a Steiger test post-MR analysis to confirm the causal direction^57^, and (4) conducted reverse MR analysis.

**To ensure the robustness of our results**, we employed several sensitivity analysis methods: (1) used different thresholds (P < 5 × 10^−8^ and P < 1 × 10^−5^) to extract IVs and calculated results accordingly, (2) utilized one primary and eight secondary analysis methods for robustness, (3) applied Cochran’s Q statistic to assess heterogeneity in IVW estimates, (4) evaluated horizontal pleiotropy using the p-value for the intercept in MR-Egger^52^ and the global test in MR-PRESSO^58^, (5) excluded outlier SNPs identified by MR-PRESSO and repeated the analysis with the remaining SNPs, and (6) used the leave-one-out method to determine if results were driven by any single SNP.

#### 4.2 Multivariable MR

We adjusted for six confounding factors in the bidirectional MVMR analysis. The IVW, Lasso, Weighted median, and MR-Egger methods estimated the causal effect. Pleiotropy was tested by comparing the MR-Egger intercept to zero. IVW was the primary method for estimating MVMR effects in the absence of pleiotropy. If pleiotropy was present, we used multivariable MR-Egger for estimation.

#### 4.3 Network MR

We used Network MR to explore indirect effects in the bidirectional causal pathways between ADHD and RLS. This involved two procedures. First, MR assessed the effect of exposure on candidate mediators (β1). Then, mediators significantly associated with exposures were used as exposures to infer causalities between mediators and the outcome (β2). The magnitude of the mediating effect was evaluated using the product of coefficients method (β1×β2), ensuring the direction of the mediating effect aligned with the overall effect. The proportion of the mediating effect was calculated by dividing the mediating effect by the overall effect. The standard error of the mediating effect was derived using the delta method^59^. For each procedure’s results, we conducted heterogeneity and horizontal pleiotropy analyses as sensitivity checks.

#### 4.4 Evaluation and correction for the bias from sample overlap

We used the UVMR method to explore the causal relationship between ADHD and RLS, with both datasets containing participants from the deCODE cohort. The maximum sample overlap rate was 2.38% (11,448 / 480,982)^60^. When evaluating the causal relationships between eight iron-sensitive imaging features, nine DTI features and RLS, the maximum sample overlap rates were 6.25% (30,056 / 480,982) and 7.61% (36,642 / 480,982), respectively, due to probable common participants from the UK Biobank. Since these overlaps were relatively small, the resulting bias was deemed negligible^61–63^. However, when exploring the relationship between ADHD and the age of first birth, the maximum sample overlap rate was 25.42% (137,993 / 542,901) due to participants from the deCODE cohort. Additionally, when exploring the causality between the age of first birth and RLS, the maximum overlap rate was 32.86% ([11,448+166,944] / 542,901) due to participants from both the deCODE cohort and the UK Biobank. To ensure robust results, we used the MRlap method^64,65^ to correct for bias from sample overlap in these analyses.

All MR analyses were conducted using R version 4.3.1, with the TwoSampleMR (0.5.7), MR-PRESSO, and MRlap (0.0.3) packages.

## Results

### 1 Stage one: exploring shared genetic architecture

The LDSC revealed a significant positive genetic correlation between ADHD and RLS (rg = 0.285, p = 9.19E-07). The HDL method produced similar results (rg = 0.271, P = 1.51E-05, **Table 1**).

**Table1.** Genome-wide genetic correlation between ADHD and RLS using LDSC and HDL.

| Diseases | LDSC |  |  |  | HDL |  |  |  |
| --- | --- | --- | --- | --- | --- | --- | --- | --- |
|  | h2 | Rg | se | P | h2 | Rg | se | P |
| ADHD | 0.086 | 0.285 | 0.058 | 9.19E-07 | 0.103 | 0.271 | 0.063 | 1.51E-05 |
| RLS | 0.007 |  |  |  | 0.006 |  |  |  |

Abbreviations: ADHD Attention deficit hyperactivity disorder, HDL High-definition likelihood method, h2 heritability, LDSC Linkage disequilibrium score regression, Rg genetic correlation estimate, RLS Restless legs syndrome, se standard error.

### 2 Stage two: exploring genetic pleiotropy

#### 2.1 Pleiotropic SNPs and risk loci

PLACO was applied to identify pleiotropic variants between ADHD and RLS at SNP-level (**Table S2**). The Manhattan plot for the result is shown in **Figure 2A**. The QQ plots demonstrated no premature divergence between observed and expected values, ruling out group stratification (**Figure 2B**). Based on PLACO results, we further identified five pleiotropic genetic loci using FUMA platform (**Table 2**). Colocalization analysis revealed a shared causal genetic variant, rs12336113 (PP.H4 = 0.78), between ADHD and RLS at locus 4. This variant is located in the intronic region of the PTPRD gene. Further tissue enrichment analysis identified that these pleiotropic SNPs are primarily enriched in various brain tissues **(Figure 2C)**.

**Figure 2.**
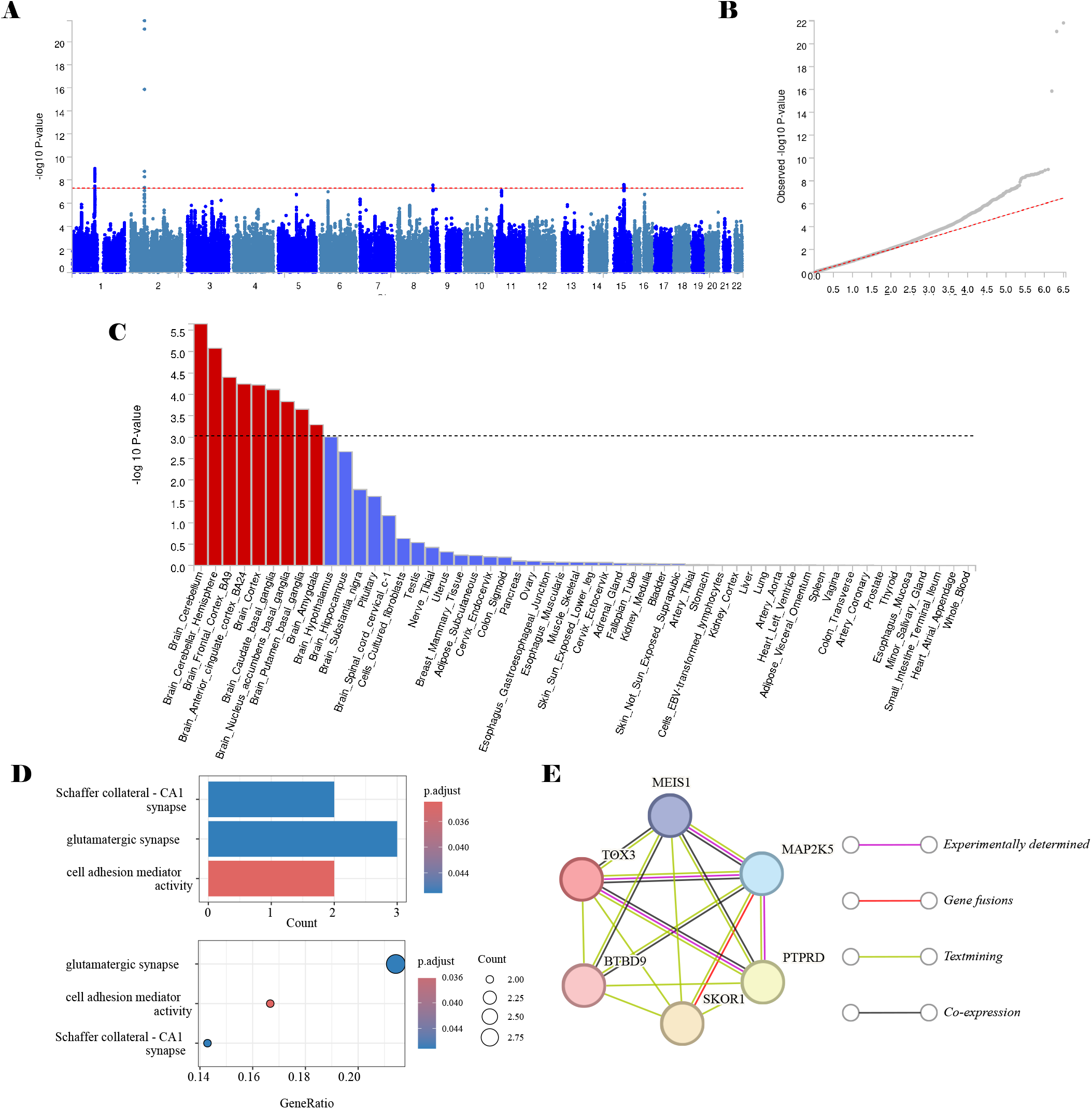
shows the results of the genetic pleiotropy analysis in stage two. A-B: The Manhattan plot and Q-Q plot based on the results of PLACO. C: Tissue enrichment results based on pleiotropic SNPs. D-E: Functional enrichment and PPI analysis results based on pleiotropic genes.

**Table 2.** Colocalized loci identified by PLACO and colocalization analysis between ADHD and RLS.

| Locus | Top SNP | Locus boundary | Effect allele | Other allele | P_PLACO | Nearest gene | Function of top SNP | PP.H4 |
| --- | --- | --- | --- | --- | --- | --- | --- | --- |
| 1 | rs12049261 | 1:107180648-107401497 | C | G | 9.99E-10 | RP11-478L17.1 | Intergenic | 9.16E-03 |
| 2 | rs11679120 | 2:66728627-66816983 | A | G | 1.50E-22 | MEIS1 | Intronic | 6.13E-02 |
| 3 | rs10815977 | 9:8858688-8865153 | A | G | 2.72E-08 | PTPRD:RP11-75C9.1 | ncRNA_exonic | 1.66E-02 |
| 4 | rs12336113 | 9:9155721-9161855 | A | G | 5.00E-08 | PTPRD | Intronic | 7.82E-01 |
| 5 | rs4275804 | 15:67687968-68124665 | C | T | 2.47E-08 | IQCH | Intronic | 7.17E-02 |

Locus boundary of each pleiotropic genomic risk locus was denoted as “chromosome: start-end” defined by FUMA for the corresponding trait pair. Abbreviations: PP.H4, Posterior Probability of Hypothesis 4

#### 2.2 Pleiotropic genes

We used MAGMA gene analysis and gene-mapping methods to translate identified SNP-level signals into gene-level signals, identifying 14 significant pleiotropic genes between ADHD and RLS (**Table S3**). GO analysis of these genes revealed enrichment in functions related to the Schaffer collateral-CA1 synapse, glutamatergic synapse, and cell adhesion mediator activity (**Figure 2D**, **Table S4**). However, KEGG analysis was not significant after FDR correction. To further elucidate the relationships among these genes, we conducted PPI analysis, which revealed that six genes closely related to neuropsychiatric disorders (MEIS1, TOX3, BTBD9, SKOR1, PTPRD, MAP2K5) form an interaction network (**Figure 2E**).

### 3 Stage three: exploring gene-driven causality

#### 3.1 Univariable MR

Using a threshold of P < 5 × 10^−8^ for selecting IVs (**Table S6**), the results indicated that ADHD increases the risk of RLS (IVW: Odds ratio (OR) = 1.203, 95% confidence interval (CI) = 1.058 to 1.368, P = 4.80 × 10^−3^, **Figure 3A**). Eight other MR methods produced consistent estimates, with six reaching statistical significance. The reverse MR analysis showed that RLS also increases the risk of ADHD (IVW: OR = 1.039, 95% CI = 1.002 to 1.077, P = 3.92 × 10^−2^, **Figure 3A**), with consistent results from eight other MR methods, six of which were statistically significant. Sensitivity analyses suggested no heterogeneity or horizontal pleiotropy interference, and leave-one-out analysis confirmed that results were not driven by any single IV. The causal direction from exposure to outcome was validated by the Steiger test. Detailed sensitivity analysis results are in **Table S7, 8**. Additionally, when extracting IVs with more lenient criteria (P = 1 × 10^−5^), the estimates continued to support the bidirectional causal relationship between ADHD and RLS (**Figure 3B**).

**Figure 3.**
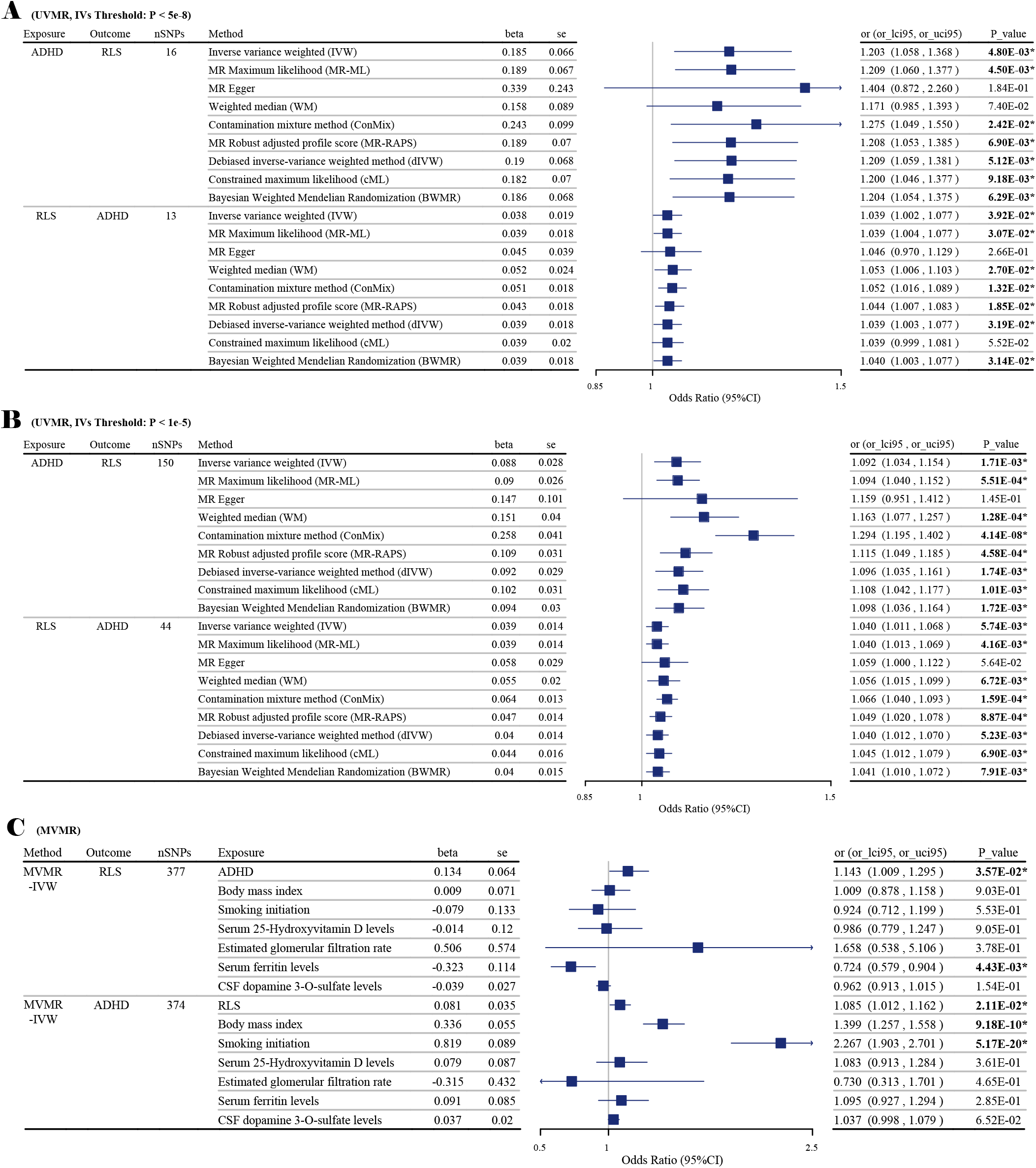
shows the results of the gene-driven causality analysis. A: Results from the UVMR analysis using a threshold of 5× 10^−8^ for extracting IVs. B: Results from the UVMR analysis using a threshold of 1× 10^−5^ for extracting IVs. C: Results from the MVMR analysis.

#### 3.2 Multivariable MR

After adjusting for BMI, smoking initiation, serum 25-hydroxyvitamin D levels, estimated glomerular filtration rate, serum ferritin levels, and CSF dopamine 3-O-sulfate levels, ADHD remained an independent risk factor for RLS (MVMR_IVW: OR = 1.143, 95% CI = 1.009 to 1.295, P = 3.57 × 10^−2^). Similarly, the effect of RLS on ADHD remained robust (MVMR_IVW: OR = 1.085, 95% CI = 1.012 to 1.162, P = 2.11 × 10^−2^). The multivariable MR-Egger intercept term was close to zero, indicating no horizontal pleiotropy. However, the heterogeneity test indicated heterogeneity across all IVs, possibly due to the inclusion of too many IVs in the MVMR analysis (**Table S9**).

#### 3 Network MR

To explore the indirect causal pathways between ADHD and RLS, we conducted Network MR analysis. For the specific screening process of mediators, please refer to **Tables S11,12**. We finally found that ADHD can indirectly increase the risk of RLS by advancing the age at first birth (indirect effect =0.021, 95% CI = 0.001 to 0.042, P = 3.76 × 10^−2^, proportion = 11.62%, **Figure 4A, C**). Considering the sample overlap between exposure and mediator (25.42% [137,993/542,901]), and between mediator and outcome (32.86% [166,944/542,901 + 11,448/542,901]), we used the MRlap method to correct for bias from sample overlap and re-estimated the causal effect. The mediating effect remained significant (indirect effect = 0.021, 95% CI = 0.001 to 0.040, P = 2.36 × 10^−2^, proportion = 4.13%).

**Figure 4.**
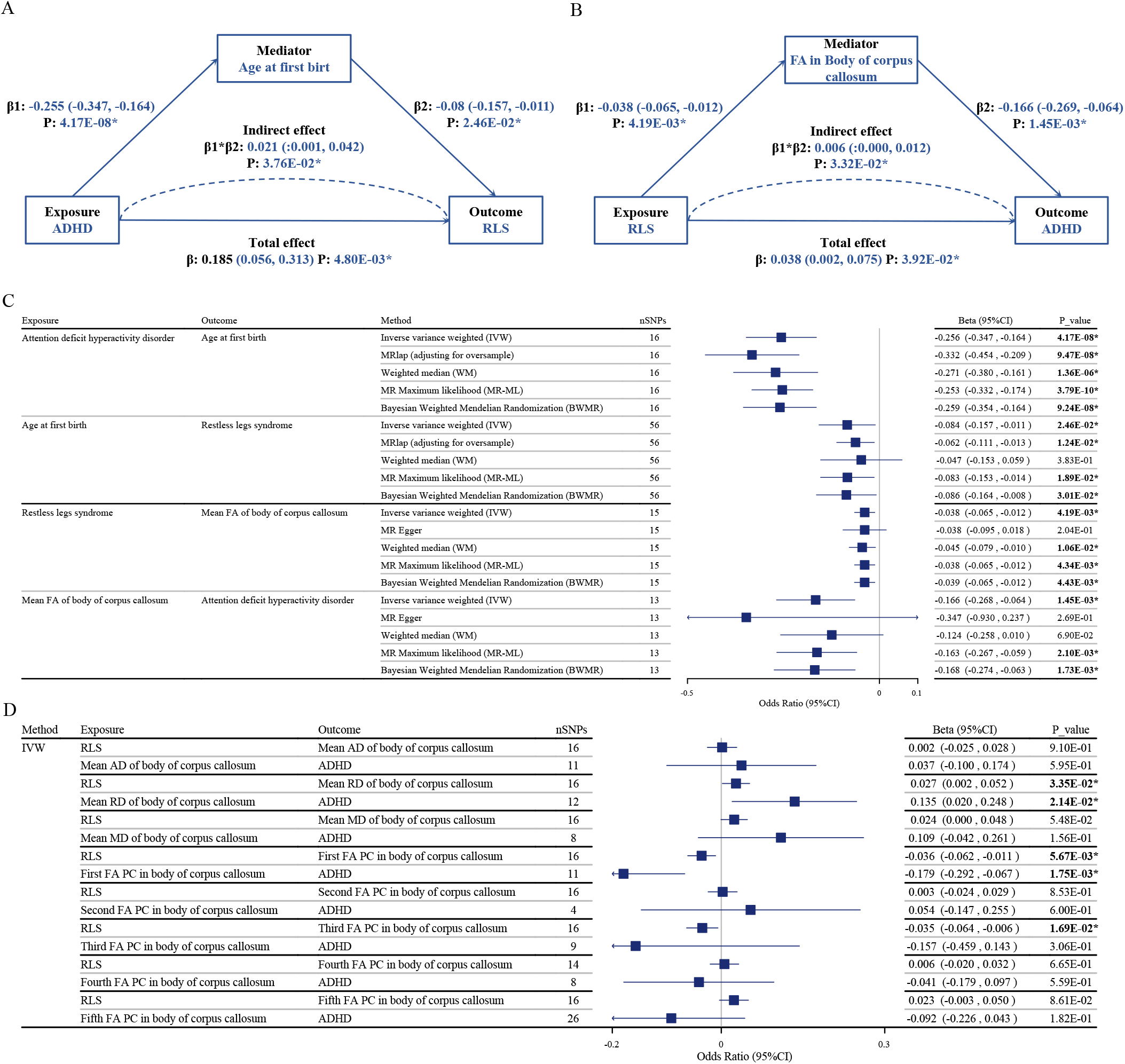
shows the bidirectional, indirect causal pathways between ADHD and RLS. A: Indirect causal pathway between ADHD, age at first birth, and RLS. B: Indirect causal pathway between RLS, FA in the body of the corpus callosum, and ADHD. C: Forest plot illustrating the relationships between exposure, mediator, and outcome. D: Forest plot displaying the potential mediating effects of various PCs of FA, as well as AD, RD, and MD, in the body of the corpus callosum. PC Principal component, FA Fractional anisotropy, MD Mean diffusivity, AD Axial diffusivity, RD Radial diffusivity.

We also found that RLS can indirectly lead to ADHD by reducing fractional anisotropy (FA) in the body of the corpus callosum (indirect effect =0.006, 95% CI = 0.000 to 0.012, P = 3.32 × 10^−2^, proportion = 16.67%, **Figure 4B, C**). To further analyze the specific changes in body of corpus callosum, we divided it into five principal components (PCs) and included them in the Network MR analysis. We found that RLS can indirectly lead to ADHD by reducing the first FA PC in the body of the corpus callosum (indirect effect =0.006, 95% CI = 0.000 to 0.013, P = 3.82 × 10^−2^). Additionally, we included the axial diffusivity (AD), radial diffusivity (RD), and mean diffusivity (MD) values of the body of the corpus callosum in the Network MR analysis and found that RLS increases the mean RD in the body of the corpus callosum (IVW: Beta = 0.027, 95% CI = 0.002 to 0.052, P = 3.35 × 10^−2^), which in turn promotes the occurrence of ADHD (IVW: Beta = 0.135, 95% CI = 0.020 to 0.248, P = 2.14 × 10^−2^, **Figure 4D**). These analyses showed no heterogeneity or horizontal pleiotropy in the MR results between exposure and mediator, and between mediator and outcome (**Table S13**).

## Discussion

This study investigates the genetic mechanisms underlying the comorbidity of ADHD and RLS, divided into three stages. In stage one, a genome-wide genetic correlation analysis revealed a shared genetic structure between the two diseases. Considering this could result from genetic pleiotropy and/or causality, we conducted further analyses. In stage two, comprehensive analyses identified pleiotropic genetic variants, risk loci, shared genes, and biological pathways. In stage three, MR analysis demonstrated a bidirectional causal relationship between ADHD and RLS. ADHD can indirectly promote the onset of RLS by advancing the age at first birth. Conversely, RLS can indirectly contribute to the onset of ADHD by reducing FA in the body of the corpus callosum, with the most significant changes in PC 1. In this process, the increase in RD, rather than the decrease in AD, plays a crucial role.

### 1 Genetic pleiotropy

Previous GWAS studies did not find significant associations between SNPs in RLS risk genes and ADHD, possibly due to a small sample size (sample size = 386) and the limited number of target genes (only MEIS1, BTBD9, and MAP2K5). Our research is based on the latest GWAS data and analysis methods. Through cross-trait PLACO analysis, we identified five shared genetic risk loci between ADHD and RLS. Colocalization analysis indicated that one of these loci (locus 4, PP.H4 = 0.78) contains a shared causal variant, rs12336113, located in an intron of the PTPRD gene. Subsequently, using Gene-mapping and MAGMA methods, we identified 14 potential shared genes between these two disorders. PPI analysis showed that six of these genes—PTPRD, MEIS1, MAP2K5, SKOR1, BTBD9, and TOX3—are closely connected.

The PTPRD gene encodes the protein tyrosine phosphatase receptor type delta, which is highly expressed in brain tissue and is associated with spatial learning, synaptic plasticity, and motor neuron axon guidance^66–68^. A study on the role of structural variation in ADHD (including 335 ADHD patients and their parents, and 2026 healthy controls) found four independent deletions within the PTPRD gene^69^, further validated by quantitative PCR. Given that PTPRD has been identified as a candidate gene for RLS^70^, and two out of four ADHD probands with PTPRD deletions reported RLS symptoms, this study suggests a possible link between PTPRD and the comorbidity of ADHD and RLS. Our findings further support this hypothesis. Additionally, the shared genetic variant rs12336113, located in an intron of PTPRD, may contribute to both ADHD and RLS by regulating PTPRD expression.

Furthermore, we identified five genes closely related to PTPRD: MEIS1, MAP2K5, SKOR1, BTBD9, and TOX3. These genes are highly expressed in brain tissue and are associated with embryonic neuronal development, iron metabolism, dopamine synthesis, and the pathogenesis of RLS^37^. The interactions and regulatory relationships of these genes with PTPRD, as well as their roles in the comorbidity of ADHD and RLS, require further investigation.

### 2 Gene-driven causality

A large cross-sectional study of Danish blood donors (who are generally required to be healthy, particularly with normal iron levels) with a sample size of 25,336 found a strong association between ADHD and RLS even after adjusting for BMI and smoking factors and excluding the interference of iron metabolism disorders^71^. This suggests that, in addition to shared pathogenesis, there may be a causal relationship between these two conditions. MR analysis uses SNPs as instrumental variables for causal inference. Since the allocation of SNPs is entirely random, unaffected by confounding factors, and occurs before the outcome, MR results are less prone to confounding bias and reverse causation compared to observational studies.

#### 2.1 ADHD, age at first birth, and RLS

As previously mentioned, numerous studies have shown a significantly higher prevalence of RLS in patients with ADHD, as well as a higher incidence of RLS in the parents of children with ADHD. Through MR analysis, we identified ADHD as an independent risk factor for RLS. This finding is supported by various models, passed the Steiger test for directionality, and is robust to heterogeneity, horizontal pleiotropy, and outliers tests. It remains consistent under different selection criteria for instrumental variables. Even after adjusting for BMI, smoking initiation, serum 25-hydroxyvitamin D levels, estimated glomerular filtration rate, serum ferritin levels, and CSF dopamine 3-O-sulfate levels in multivariable MR analysis, the association remains, further enhancing the credibility of our conclusion. Additionally, network MR analysis revealed that ADHD might indirectly promote RLS onset by advancing the age at first birth.

Previous cross-sectional, cohort, and genetic studies support that ADHD is an independent risk factor for early childbearing, possibly due to increased libido and risk-taking behavior leading to unintended pregnancies^72–75^. Previous research also supports pregnancy history as an independent risk factor for RLS^76,77^. Women without a history of pregnancy have RLS incidence rates similar to men, while women with a pregnancy history have significantly higher rates of RLS^78^. Furthermore, the risk of future RLS increases with each subsequent pregnancy^79^. This may be due to long-term effects of iron deficiency, vitamin D deficiency, hormonal changes, and weight gain during pregnancy^80^. Thus, women who have children at a younger age may be exposed to the risks associated with pregnancy for a longer period, leading to a cumulative effect that ultimately increases the incidence of RLS. From another perspective, early childbearing is closely related to adverse maternal health outcomes. Studies have shown that early childbearing increases the risk of obesity, diabetes, and COPD in mothers^81–84^, which are significant risk factors for RLS^31,85–87^.

Therefore, we believe that ADHD may increase the risk of RLS by advancing the age at first birth. However, it should be noted that this explanation only addresses the indirect causal pathway for female ADHD patients developing RLS. Whether this pathway applies to males requires further research. Additionally, the relationship between age at first birth and RLS needs more validation from observational studies.

#### 2.2 RLS, demyelination of the corpus callosum body, and ADHD

Over a long-term follow-up, a large cohort study focusing on Australian children (sample size = 2120)^88^ and another on 7072 Chinese adolescents^89^ both support RLS as an independent risk factor for future ADHD. Through MR analysis, we also found that RLS can promote the onset of ADHD. This conclusion was supported by various sensitivity analyses and remained robust after adjusting for multiple confounding factors in multivariable MR analysis.

Patients with ADHD and RLS may both exhibit changes in white matter tracts. Previous DTI studies found reduced FA values in the corpus callosum of RLS patients^90^. Byeong-Yeul Lee and his team identified a reduction in the body of the corpus callosum through thickness analysis^91^, while Deniz Sigirli and his team found significant changes in the posterior midbody of the corpus callosum through shape analysis^92^. A large meta-analysis encompassing 32 datasets (26 involving children and 6 involving adults) showed that one of the most significant and consistent white matter changes in ADHD patients is the reduced FA value in the body of the corpus callosum^93^. These studies suggest that changes in the corpus callosum body fibers are a common pathological feature of both diseases. Network MR analysis further revealed that RLS can indirectly contribute to the onset of ADHD by reducing FA in the body of the corpus callosum, with the most significant changes in PC 1. In this process, the increase in RD, rather than the decrease in AD, plays a crucial role.

FA, AD, and RD are common parameters in DTI. FA reflects the degree of water molecule diffusion anisotropy; its reduction indicates disruption in the integrity and organization of white matter fibers. RD reflects the diffusion of water molecules perpendicular to the main fiber direction; its increase is mainly seen in demyelination and less frequently in axonal damage. AD indicates the diffusion of water molecules parallel to the main fiber direction; its reduction primarily reflects axonal damage. Therefore, reduced FA and increased RD in the body of the corpus callosum, without significant changes in AD, suggest demyelination in this region. Animal and clinical studies have shown that iron metabolism abnormalities can cause brain demyelination^94–97^. A study examining postmortem brain tissue samples of RLS patients (n=11) and healthy controls (n=11) using WB found less myelin and loss of myelin integrity in RLS brains, coupled with decreased ferritin and transferrin in the myelin fractions^98^. The body of the corpus callosum comprises highly myelinated fibers that interconnect the two cerebral hemispheres, playing a crucial role in sensory integration, motor coordination^99^, and emotional-cognitive regulation^100^. Our study suggests that demyelination in the corpus callosum body of RLS patients may indirectly lead to the development of ADHD. The specific relationship between demyelination, iron metabolism disorders, and these two diseases is an intriguing area for future research.

### 3 Limitations

Our study has the following limitations: (1) The GWAS data used in our analysis are derived from European populations, limiting the generalizability of our conclusions. (2) Observational studies indicate that the association between different ADHD subtypes and RLS varies in strength, which needs further clarification through stratified GWAS analyses in the future. (3) Some MR analyses may be affected by mild sample overlap. We have rigorously assessed the overlap rate and corrected the results using relevant algorithms (MRlap) when necessary. (4) Dopaminergic system dysfunction may be a common pathological change between ADHD and RLS. However, in the GWAS database, we currently lack a better marker than CSF dopamine 3-O-sulfate levels to describe this change. Thus, when using MR analysis to explore the causal relationship between ADHD and RLS, we cannot entirely rule out the possibility of pleiotropic IVs. Nevertheless, considering we employed multiple models robust to horizontal pleiotropy and the results remained stable, this issue is not overly concerning. (5) From a triangulation of evidence perspective, we urge future observational studies to further verify the following MR analysis results: A. Explore whether ADHD can be an independent risk factor for RLS in **cohort studies**; B. Determine whether early childbearing is an independent risk factor for RLS. (6) We also call for future research to further elucidate the potential roles of the PTPRD gene, iron metabolism disorders, and demyelination of the corpus callosum body in the comorbidity of ADHD and RLS.

## Conclusions

ADHD and RLS share a common genetic structure, with PTPRD being the most strongly supported pleiotropic gene. The potential roles of other genes require further investigation. There is a gene-driven bidirectional causal relationship between ADHD and RLS. ADHD may indirectly promote the onset of RLS by advancing the age of first birth, while RLS may indirectly contribute to the development of ADHD by causing demyelination in the body of the corpus callosum.

## Data Availability

All data produced in the present study are available upon reasonable request to the authors.

## References

1. Astle DE, Holmes J, Kievit R, Gathercole SE. Annual Research Review: The transdiagnostic revolution in neurodevelopmental disorders. Journal of child psychology and psychiatry, and allied disciplines. 2022; 63 (4): 397–417.

2. Global, regional, and national burden of 12 mental disorders in 204 countries and territories, 1990-2019: a systematic analysis for the Global Burden of Disease Study 2019. The lancet Psychiatry. 2022; 9 (2): 137–150.

3. Thomas R, Sanders S, Doust J, Beller E, Glasziou P. Prevalence of attention-deficit/hyperactivity disorder: a systematic review and meta-analysis. Pediatrics. 2015; 135 (4): e994–1001.

4. Sibley MH, Mitchell JT, Becker SP. Method of adult diagnosis influences estimated persistence of childhood ADHD: a systematic review of longitudinal studies. The lancet Psychiatry. 2016; 3 (12): 1157–1165.

5. Doshi JA, Hodgkins P, Kahle J, et al. Economic impact of childhood and adult attention-deficit/hyperactivity disorder in the United States. Journal of the American Academy of Child and Adolescent Psychiatry. 2012; 51 (10): 990–1002.e1002.

6. Berger K, Kurth T. RLS epidemiology--frequencies, risk factors and methods in population studies. Movement disorders : official journal of the Movement Disorder Society. 2007; 22 Suppl 18: S420–423.

7. Allen RP, Picchietti D, Hening WA, Trenkwalder C, Walters AS, Montplaisi J. Restless legs syndrome: diagnostic criteria, special considerations, and epidemiology. A report from the restless legs syndrome diagnosis and epidemiology workshop at the National Institutes of Health. Sleep medicine. 2003; 4 (2): 101–119.

8. Chervin RD, Archbold KH, Dillon JE, et al. Associations between symptoms of inattention, hyperactivity, restless legs, and periodic leg movements. Sleep. 2002; 25 (2): 213–218.

9. Schredl M, Alm B, Sobanski E. Sleep quality in adult patients with attention deficit hyperactivity disorder (ADHD). European archives of psychiatry and clinical neuroscience. 2007; 257 (3): 164–168.

10. Chervin RD, Dillon JE, Bassetti C, Ganoczy DA, Pituch KJ. Symptoms of Sleep Disorders, Inattention, and Hyperactivity in Children. Sleep. 1997; 20 (12): 1185–1192.

11. Furudate N, Komada Y, Kobayashi M, Nakajima S, Inoue Y. Daytime dysfunction in children with restless legs syndrome. Journal of the neurological sciences. 2014; 336 (1-2): 232–236.

12. Konofal E, Cortese S, Marchand M, Mouren MC, Arnulf I, Lecendreux M. Impact of restless legs syndrome and iron deficiency on attention-deficit/hyperactivity disorder in children. Sleep medicine. 2007; 8 (7-8): 711–715.

13. Wagner ML, Walters AS, Fisher BC. Symptoms of attention-deficit/hyperactivity disorder in adults with restless legs syndrome. Sleep. 2004; 27 (8): 1499–1504.

14. Picchietti DL, Underwood DJ, Farris WA, et al. Further studies on periodic limb movement disorder and restless legs syndrome in children with attention-deficit hyperactivity disorder. Movement disorders : official journal of the Movement Disorder Society. 1999; 14 (6): 1000–1007.

15. Gao X, Lyall K, Palacios N, Walters AS, Ascherio A. RLS in middle aged women and attention deficit/hyperactivity disorder in their offspring. Sleep medicine. 2011; 12 (1): 89–91.

16. Cortese S, Konofal E, Lecendreux M, et al. Restless legs syndrome and attention-deficit/hyperactivity disorder: a review of the literature. Sleep. 2005; 28 (8): 1007–1013.

17. Konofal E, Lecendreux M, Arnulf I, Mouren MC. Iron deficiency in children with attention-deficit/hyperactivity disorder. Archives of pediatrics & adolescent medicine. 2004; 158 (12): 1113–1115.

18. Allen RP, Barker PB, Wehrl FW, Song HK, Earley CJ. MRI measurement of brain iron in patients with restless legs syndrome. Neurology. 2001; 56 (2): 263–265.

19. Biederman J, Faraone SV. Attention-deficit hyperactivity disorder. Lancet (London, England). 2005; 366 (9481): 237–248.

20. Gamaldo CE, Benbrook AR, Allen RP, Scott JA, Henning WA, Earley CJ. Childhood and adult factors associated with restless legs syndrome (RLS) diagnosis. Sleep medicine. 2007; 8 (7-8): 716–722.

21. Tseng PT, Cheng YS, Yen CF, et al. Peripheral iron levels in children with attention-deficit hyperactivity disorder: a systematic review and meta-analysis. Scientific reports. 2018; 8 (1): 788.

22. Bener A, Kamal M, Bener H, Bhugra D. Higher prevalence of iron deficiency as strong predictor of attention deficit hyperactivity disorder in children. Annals of medical and health sciences research. 2014; 4 (Suppl 3): S291–297.

23. Erikson KM, Jones BC, Hess EJ, Zhang Q, Beard JL. Iron deficiency decreases dopamine D1 and D2 receptors in rat brain. Pharmacology, biochemistry, and behavior. 2001; 69 (3-4): 409–418.

24. Bianco LE, Wiesinger J, Earley CJ, Jones BC, Beard JL. Iron deficiency alters dopamine uptake and response to L-DOPA injection in Sprague-Dawley rats. Journal of neurochemistry. 2008; 106 (1): 205–215.

25. Schimmelmann BG, Friedel S, Nguyen TT, et al. Exploring the genetic link between RLS and ADHD. Journal of psychiatric research. 2009; 43 (10): 941–945.

26. Kurilshikov A, Medina-Gomez C, Bacigalupe R, et al. Large-scale association analyses identify host factors influencing human gut microbiome composition. Nature genetics. 2021; 53 (2): 156-165.

27. Demontis D, Walters GB, Athanasiadis G, et al. Genome-wide analyses of ADHD identify 27 risk loci, refine the genetic architecture and implicate several cognitive domains. Nature genetics. 2023; 55 (2): 198–208.

28. Demontis D, Walters RK, Martin J, et al. Discovery of the first genome-wide significant risk loci for attention deficit/hyperactivity disorder. Nature genetics. 2019; 51 (1): 63–75.

29. Didriksen M, Nawaz MS, Dowsett J, et al. Large genome-wide association study identifies three novel risk variants for restless legs syndrome. Communications biology. 2020; 3 (1): 703.

30. Hagman E, Danielsson P, Brandt L, Svensson V, Ekbom A, Marcus C. Childhood Obesity, Obesity Treatment Outcome, and Achieved Education: A Prospective Cohort Study. The Journal of adolescent health : official publication of the Society for Adolescent Medicine. 2017; 61 (4): 508–513.

31. Lin S, Zhang H, Gao T, et al. The association between obesity and restless legs syndrome: A systemic review and meta-analysis of observational studies. Journal of affective disorders. 2018; 235: 384–391.

32. Huang A, Wu K, Cai Z, Lin Y, Zhang X, Huang Y. Association between postnatal second-hand smoke exposure and ADHD in children: a systematic review and meta-analysis. Environmental science and pollution research international. 2021; 28 (2): 1370–1380.

33. Lee SS, Humphreys KL, Flory K, Liu R, Glass K. Prospective association of childhood attention-deficit/hyperactivity disorder (ADHD) and substance use and abuse/dependence: a meta-analytic review. Clinical psychology review. 2011; 31 (3): 328–341.

34. Sucksdorff M, Brown AS, Chudal R, et al. Maternal Vitamin D Levels and the Risk of Offspring Attention-Deficit/Hyperactivity Disorder. Journal of the American Academy of Child and Adolescent Psychiatry. 2021; 60 (1): 142–151.e142.

35. Balaban H, Yıldız Ö K, Çil G, et al. Serum 25-hydroxyvitamin D levels in restless legs syndrome patients. Sleep medicine. 2012; 13 (7): 953–957.

36. Simões ESAC, Miranda AS, Rocha NP, Teixeira AL. Neuropsychiatric Disorders in Chronic Kidney Disease. Frontiers in pharmacology. 2019; 10: 932.

37. Guo S, Huang J, Jiang H, et al. Restless Legs Syndrome: From Pathophysiology to Clinical Diagnosis and Management. Frontiers in aging neuroscience. 2017; 9: 171.

38. Salas RE, Gamaldo CE, Allen RP. Update in restless legs syndrome. Current opinion in neurology. 2010; 23 (4): 401–406.

39. Bulik-Sullivan B, Finucane HK, Anttila V, et al. An atlas of genetic correlations across human diseases and traits. Nature genetics. 2015; 47 (11): 1236–1241.

40. Auton A, Brooks LD, Durbin RM, et al. A global reference for human genetic variation. Nature. 2015; 526 (7571): 68–74.

41. Ning Z, Pawitan Y, Shen X. High-definition likelihood inference of genetic correlations across human complex traits. Nature genetics. 2020; 52 (8): 859–864.

42. Ray D, Chatterjee N. A powerful method for pleiotropic analysis under composite null hypothesis identifies novel shared loci between Type 2 Diabetes and Prostate Cancer. PLoS genetics. 2020; 16 (12): e1009218.

43. Watanabe K, Taskesen E, van Bochoven A, Posthuma D. Functional mapping and annotation of genetic associations with FUMA. Nature communications. 2017; 8 (1): 1826.

44. Giambartolomei C, Vukcevic D, Schadt EE, et al. Bayesian test for colocalisation between pairs of genetic association studies using summary statistics. PLoS genetics. 2014; 10 (5): e1004383.

45. Ogata H, Goto S, Sato K, Fujibuchi W, Bono H, Kanehisa M. KEGG: Kyoto Encyclopedia of Genes and Genomes. Nucleic acids research. 1999; 27 (1): 29–34.

46. Harris MA, Clark J, Ireland A, et al. The Gene Ontology (GO) database and informatics resource. Nucleic acids research. 2004; 32 (Database issue): D258–261.

47. Szklarczyk D, Kirsch R, Koutrouli M, et al. The STRING database in 2023: protein-protein association networks and functional enrichment analyses for any sequenced genome of interest. Nucleic acids research. 2023; 51 (D1): D638–d646.

48. Skrivankova VW, Richmond RC, Woolf BAR, et al. Strengthening the reporting of observational studies in epidemiology using mendelian randomisation (STROBE-MR): explanation and elaboration. BMJ (Clinical research ed). 2021; 375: n2233.

49. Burgess S, Bowden J, Fall T, Ingelsson E, Thompson SG. Sensitivity Analyses for Robust Causal Inference from Mendelian Randomization Analyses with Multiple Genetic Variants. Epidemiology (Cambridge, Mass). 2017; 28 (1): 30–42.

50. Xue H, Shen X, Pan W. Constrained maximum likelihood-based Mendelian randomization robust to both correlated and uncorrelated pleiotropic effects. American journal of human genetics. 2021; 108 (7): 1251–1269.

51. Bowden J, Davey Smith G, Haycock PC, Burgess S. Consistent Estimation in Mendelian Randomization with Some Invalid Instruments Using a Weighted Median Estimator. Genetic epidemiology. 2016; 40 (4): 304–314.

52. Bowden J, Davey Smith G, Burgess S. Mendelian randomization with invalid instruments: effect estimation and bias detection through Egger regression. International journal of epidemiology. 2015; 44 (2): 512–525.

53. Burgess S, Foley CN, Allara E, Staley JR, Howson JMM. A robust and efficient method for Mendelian randomization with hundreds of genetic variants. Nature communications. 2020; 11 (1): 376.

54. Yu K, Chen XF, Guo J, et al. Assessment of bidirectional relationships between brain imaging-derived phenotypes and stroke: a Mendelian randomization study. BMC medicine. 2023; 21 (1): 271.

55. Zhao J, Ming J, Hu X, Chen G, Liu J, Yang C. Bayesian weighted Mendelian randomization for causal inference based on summary statistics. Bioinformatics (Oxford, England). 2020; 36 (5): 1501–1508.

56. Park S. A Causal and Inverse Relationship between Plant-Based Diet Intake and in a Two-Sample Mendelian Randomization Study. Foods. 2023; 12 (3).

57. Hemani G, Tilling K, Davey Smith G. Orienting the causal relationship between imprecisely measured traits using GWAS summary data. PLoS genetics. 2017; 13 (11): e1007081.

58. Verbanck M, Chen CY, Neale B, Do R. Detection of widespread horizontal pleiotropy in causal relationships inferred from Mendelian randomization between complex traits and diseases. Nature genetics. 2018; 50 (5): 693–698.

59. Carter AR, Gill D, Davies NM, et al. Understanding the consequences of education inequality on cardiovascular disease: mendelian randomisation study. BMJ (Clinical research ed). 2019; 365: l1855.

60. Burgess S, Davies NM, Thompson SG. Bias due to participant overlap in two-sample Mendelian randomization. Genetic epidemiology. 2016; 40 (7): 597–608.

61. Niu PP, Wang X, Xu YM. Causal effects of serum testosterone levels on brain volume: a sex-stratified Mendelian randomization study. Journal of endocrinological investigation. 2023; 46 (9): 1787–1798.

62. Zhao L, Zhao W, Cao J, Tu Y. Causal relationships between migraine and microstructural white matter: a Mendelian randomization study. The journal of headache and pain. 2023; 24 (1): 10.

63. Zanoaga MD, Friligkou E, He J, et al. Brainwide Mendelian Randomization Study of Anxiety Disorders and Symptoms. Biological psychiatry. 2024; 95 (8): 810–817.

64. Mounier N, Kutalik Z. Bias correction for inverse variance weighting Mendelian randomization. Genetic epidemiology. 2023; 47 (4): 314–331.

65. Galimberti M, Levey DF, Deak JD, Zhou H, Stein MB, Gelernter J. Genetic influences and causal pathways shared between cannabis use disorder and other substance use traits. Molecular psychiatry. 2024.

66. Mizuno K, Hasegawa K, Katagiri T, Ogimoto M, Ichikawa T, Yakura H. MPTP delta, a putative murine homolog of HPTP delta, is expressed in specialized regions of the brain and in the B-cell lineage. Molecular and cellular biology. 1993; 13 (9): 5513–5523.

67. Uetani N, Chagnon MJ, Kennedy TE, Iwakura Y, Tremblay ML. Mammalian motoneuron axon targeting requires receptor protein tyrosine phosphatases sigma and delta. The Journal of neuroscience : the official journal of the Society for Neuroscience. 2006; 26 (22): 5872–5880.

68. Uetani N, Kato K, Ogura H, et al. Impaired learning with enhanced hippocampal long-term potentiation in PTPdelta-deficient mice. The EMBO journal. 2000; 19 (12): 2775–2785.

69. Elia J, Gai X, Xie HM, et al. Rare structural variants found in attention-deficit hyperactivity disorder are preferentially associated with neurodevelopmental genes. Molecular psychiatry. 2010; 15 (6): 637–646.

70. Schormair B, Kemlink D, Roeske D, et al. PTPRD (protein tyrosine phosphatase receptor type delta) is associated with restless legs syndrome. Nature genetics. 2008; 40 (8): 946–948.

71. Didriksen M, Thørner LW, Erikstrup C, et al. Self-reported restless legs syndrome and involuntary leg movements during sleep are associated with symptoms of attention deficit hyperactivity disorder. Sleep medicine. 2019; 57: 115–121.

72. Soldati L, Bianchi-Demicheli F, Schockaert P, et al. Association of ADHD and hypersexuality and paraphilias. Psychiatry research. 2021; 295: 113638.

73. Besag FM. ADHD treatment and pregnancy. Drug safety. 2014; 37 (6): 397–408.

74. Hinshaw SP, Nguyen PT, O’Grady SM, Rosenthal EA. Annual Research Review: Attention-deficit/hyperactivity disorder in girls and women: underrepresentation, longitudinal processes, and key directions. Journal of child psychology and psychiatry, and allied disciplines. 2022; 63 (4): 484–496.

75. Mills MC, Tropf FC, Brazel DM, et al. Identification of 371 genetic variants for age at first sex and birth linked to externalising behaviour. Nature human behaviour. 2021; 5 (12): 1717–1730.

76. Allen RP, Picchietti DL, Garcia-Borreguero D, et al. Restless legs syndrome/Willis-Ekbom disease diagnostic criteria: updated International Restless Legs Syndrome Study Group (IRLSSG) consensus criteria--history, rationale, description, and significance. Sleep medicine. 2014; 15 (8): 860–873.

77. Manconi M, Ulfberg J, Berger K, et al. When gender matters: restless legs syndrome. Report of the “RLS and woman” workshop endorsed by the European RLS Study Group. Sleep medicine reviews. 2012; 16 (4): 297–307.

78. Pantaleo NP, Hening WA, Allen RP, Earley CJ. Pregnancy accounts for most of the gender difference in prevalence of familial RLS. Sleep medicine. 2010; 11 (3): 310–313.

79. Berger K, Luedemann J, Trenkwalder C, John U, Kessler C. Sex and the risk of restless legs syndrome in the general population. Archives of internal medicine. 2004; 164 (2): 196–202.

80. Gupta R, Dhyani M, Kendzerska T, et al. Restless legs syndrome and pregnancy: prevalence, possible pathophysiological mechanisms and treatment. Acta neurologica Scandinavica. 2016; 133 (5): 320–329.

81. Guo HJ, Ye YL, Gao YF, Liu ZH. Age at first birth is associated with the likelihood of frailty in middle-aged and older women: A population-based analysis from NHANES 1999-2018. Maturitas. 2024; 181: 107904.

82. Lopez-Arana S, Burdorf A, Avendano M. Trends in overweight by educational level in 33 low- and middle-income countries: the role of parity, age at first birth and breastfeeding. Obesity reviews : an official journal of the International Association for the Study of Obesity. 2013; 14 (10): 806–817.

83. Pandeya N, Huxley RR, Chung HF, et al. Female reproductive history and risk of type 2 diabetes: A prospective analysis of 126 721 women. Diabetes, obesity & metabolism. 2018; 20 (9): 2103–2112.

84. Pirkle CM, de Albuquerque Sousa AC, Alvarado B, Zunzunegui MV. Early maternal age at first birth is associated with chronic diseases and poor physical performance in older age: cross-sectional analysis from the International Mobility in Aging Study. BMC public health. 2014; 14: 293.

85. Schipper SBJ, Van Veen MM, Elders PJM, et al. Sleep disorders in people with type 2 diabetes and associated health outcomes: a review of the literature. Diabetologia. 2021; 64 (11): 2367–2377.

86. Vanfleteren LE, Beghe B, Andersson A, Hansson D, Fabbri LM, Grote L. Multimorbidity in COPD, does sleep matter? European journal of internal medicine. 2020; 73: 7–15.

87. Thi Truong BE, Sung FC, Lin CL, Hang LW, Teng YK, Tzeng YL. A follow-up study on restless legs syndrome in chronic obstructive pulmonary disease population. Sleep medicine. 2021; 80: 9–15.

88. Galera C, Collet O, Orri M, et al. Prospective associations between ADHD symptoms and physical conditions from early childhood to adolescence: a population-based longitudinal study. The Lancet Child & adolescent health. 2023; 7 (12): 863–874.

89. Liu X, Liu ZZ, Liu BP, Sun SH, Jia CX. Associations between sleep problems and ADHD symptoms among adolescents: findings from the Shandong Adolescent Behavior and Health Cohort (SABHC). Sleep. 2020; 43 (6).

90. Chang Y, Paik JS, Lee HJ, et al. Altered white matter integrity in primary restless legs syndrome patients: diffusion tensor imaging study. Neurological research. 2014; 36 (8): 769–774.

91. Lee BY, Kim J, Connor JR, Podskalny GD, Ryu Y, Yang QX. Involvement of the central somatosensory system in restless legs syndrome: A neuroimaging study. Neurology. 2018; 90 (21): e1834–e1841.

92. Sigirli D, Gunes A, Turan Ozdemir S, Ercan I, Durmus Y, Erdemli Gursel B. Statistical shape analysis of corpus callosum in restless leg syndrome. Neurological research. 2020; 42 (9): 760–766.

93. Parlatini V, Itahashi T, Lee Y, et al. White matter alterations in Attention-Deficit/Hyperactivity Disorder (ADHD): a systematic review of 129 diffusion imaging studies with meta-analysis. Molecular psychiatry. 2023; 28 (10): 4098–4123.

94. Yu GS, Steinkirchner TM, Rao GA, Larkin EC. Effect of prenatal iron deficiency on myelination in rat pups. The American journal of pathology. 1986; 125 (3): 620–624.

95. Ortiz E, Pasquini JM, Thompson K, et al. Effect of manipulation of iron storage, transport, or availability on myelin composition and brain iron content in three different animal models. Journal of neuroscience research. 2004; 77 (5): 681–689.

96. Beard JL, Wiesinger JA, Connor JR. Pre- and postweaning iron deficiency alters myelination in Sprague-Dawley rats. Developmental neuroscience. 2003; 25 (5): 308–315.

97. Kim J, Wessling-Resnick M. Iron and mechanisms of emotional behavior. The Journal of nutritional biochemistry. 2014; 25 (11): 1101–1107.

98. Connor JR, Ponnuru P, Lee BY, et al. Postmortem and imaging based analyses reveal CNS decreased myelination in restless legs syndrome. Sleep medicine. 2011; 12 (6): 614–619.

99. Hofer S, Frahm J. Topography of the human corpus callosum revisited--comprehensive fiber tractography using diffusion tensor magnetic resonance imaging. NeuroImage. 2006; 32 (3): 989–994.

100. Barkley RA. A critique of current diagnostic criteria for attention deficit hyperactivity disorder: clinical and research implications. Journal of developmental and behavioral pediatrics : JDBP. 1990; 11 (6): 343–352.

